# Morphological changes after cranial fractionated photon radiotherapy: localized loss of white matter and grey matter volume with increasing dose

**DOI:** 10.1101/2021.01.04.20248933

**Authors:** SHJ Nagtegaal, S David, EE van Grinsven, MJE van Zandvoort, E Seravalli, TJ Snijders, MEP Philippens, JJC Verhoeff

**Affiliations:** Department of Radiation Oncology, University Medical Center Utrecht, The Netherlands, HP Q 00.3.11, PO box 85500, 3508 GA, Utrecht, the Netherlands; UMC Utrecht Brain Center, Department of Neurology & Neurosurgery, University Medical Center Utrecht, The Netherlands, HP L 01.310, PO box 85500, 3508 GA, Utrecht, the Netherlands

## Abstract

**Purpose:** Numerous brain MR imaging studies have been performed to understand radiation-induced cognitive decline. However, many of them focus on a single region of interest, e.g. cerebral cortex or hippocampus. In this study, we use deformation-based morphometry (DBM) and voxel-based morphometry (VBM) to measure the morphological changes in patients receiving fractionated photon RT, and relate these to the dose. Additionally, we study tissue specific volume changes in white matter (WM), grey matter (GM), cerebrospinal fluid and total intracranial volume (TIV).

**Methods and Materials:** From our database, we selected 28 patients with MRI of high quality available at baseline and 1 year after RT. Scans were rigidly registered to each other, and to the planning CT and dose file. We used DBM to study non-tissue-specific volumetric changes, and VBM to study volume loss in grey matter. Observed changes were then related to the applied radiation dose (EQD2). Additionally, brain tissue was segmented into WM, GM and cerebrospinal fluid, and changes in these volumes and TIV were tested.

**Results:** Performing DBM resulted in clusters of dose-dependent volume loss 1 year after RT seen throughout the brain. Both WM and GM were affected; within the latter both cerebral cortex and subcortical nuclei show volume loss. Volume loss rates ranging from 5.3 to 15.3%/30 Gy were seen in the cerebral cortical regions in which more than 40% of voxels were affected. In VBM, similar loss rates were seen in the cortex and nuclei. The total volume of WM and GM significantly decreased with rates of 5.8% and 2.1%, while TIV remained unchanged as expected.

**Conclusions:** Radiotherapy is associated with dose-dependent intracranial morphological changes throughout the entire brain. Therefore, we will consider to revise sparing of organs at risk based on future cognitive and neurofunctional data.

## Introduction

Radiation-induced brain injury is a phenomenon experienced after radiotherapy (RT) for brain tumors (1, 2). Anatomical and functional changes can lead to cognitive impairments, ranging from mild symptoms to severe dementia-like states, and occur in 50-90% of cases. This phenomenon is seen in patients receiving treatment for primary brain tumors, as well as those receiving whole-brain radiotherapy for brain metastases and prophylactic cranial radiotherapy.

Advances in imaging techniques have allowed the examination of the precise morphological changes in the brain after RT. Changes of white matter (WM) (3), cerebral cortex (4–7), and subcortical grey matter (GM) structures (8–10) have already been linked to received dose in several studies. However, these investigations have focused on specific parts or structures of the brain, which may limit the generalizability of the acquired knowledge. Techniques are available to analyze the brain in its entirety, in order to give a comprehensive estimation of the effect of RT on the brain.

One such technique is deformation based morphometry (DBM) (11, 12). Here, the entire brain is analyzed, and no pre-specification of tissue type or brain region is made beforehand. Pre-RT and post-RT MRI scans are non-linearly registered to the stereotactic Montreal Neurological Institute (MNI) brain template, and the transformations applied during these steps are recorded as 3D deformation fields. These fields can be used to determine the volume changes after RT, which in turn can be related to the received dose. Investigations in epilepsy and related research have successfully applied the DBM method previously (13, 14). The DBM model can be extended by adding explicit tissue segmentations. Healthy brain tissue can be segmented into WM, GM and cerebrospinal fluid (CSF). To study the GM in more detail, GM segments can be fused with 3D deformation fields in order to perform a tissue-specific investigation, often referred to as voxel-based morphometry (VBM) (15). In VBM, the changes in each voxel between pre-RT and post-RT GM maps are measured for each patient. Next to global changes in brain tissue volume, one may also study the sum of GM, WM and CSF volumes, which together make up the total intracranial volume (TIV) (16). VBM is a commonly applied tool in neuroscientific studies on aging (17, 18), while TIV is more commonly used to study brain development (19).

In this study, we use DBM and VBM to measure the morphological changes in glioma patients receiving fractionated photon RT, and relate these to the dose. Additionally, we study gross volume changes in WM, GM, CSF and TIV.

## Methods

### Patient selection and data collection

We retrospectively identified scans from patients treated with RT for grade II-IV glioma at the department of Radiation Oncology in 2016 and 2017. This specific period was chosen because all glioma patients were scanned on the same MRI scanner, using the same protocols. As the MRI protocol was updated after 2017, more recently treated patients were not included to maintain data homogeneity. Patients were eligible for inclusion when the following criteria were met: treatment planning CT and MRI present, and of sufficient resolution (see below); progression free survival of at least 270 days after RT; at least 1 follow-up MRI between 270 days and 360 days after RT present, and of sufficient resolution.

Clinical MRI and CT scans made for RT treatment planning were extracted from patient records and anonymized, along with all follow-up MRIs, and clinical and demographic characteristics. Informed consent for this retrospective study was waived by our institutional review board.

### Image acquisition

For every patient the planning CT and pre-RT MRI were collected, as well as all available follow-up MRIs. MR images were acquired on the same 3T scanner (Philips Ingenia, Philips Medical Systems, Best, The Netherlands) as part of routine clinical care. T1-weighted MR images were acquired with a 3D spoiled gradient (TFE) sequence without gadolinium enhancement with the following parameters: TR = 8.1 ms, TE = 3.7 ms, flip angle = 8°, 213 axial slices, matrix: 207 x 289, voxel resolution 0.96 × 0.96 x 1.00 mm^3^. The planning CT scans were acquired on a Brilliance Big bore scanner (Philips Medical Systems, Best, The Netherlands), with a tube potential of 120 kVp, with use of a matrix size of 512 × 512 and 0.65 × 0.65 × 3.0 mm^3^ voxel size.

Imaging was used for three different methods: DBM, VBM and analysis of global tissue volumes (total GM, WM, CSF and TIV). For each of these methods, we analyzed the difference in volumes between baseline (pre-RT) and 1 year follow-up (post-RT). The latter was defined as the time point closest to 360 days after start of RT for which an MRI was available.

### Image processing

Pre-RT and post-RT MRI scans were rigidly registered to each other, and to the planning CT and dose file. The MRI scans were processed automatically with the Computational Anatomy Toolbox (CAT12) (20). First, MR images were rigidly co-registered to each other, followed by image de-noising and segmentation into GM, WM and CSF. Then, the MR images were nonlinearly registered to standard stereotaxic MNI space (21) of 1.5 mm isotropic resolution and smoothed using an 8mm kernel size. While spatial smoothing decreases the effective spatial resolution by incorporating information from neighboring voxels, it also increases the signal to noise ratio (SNR). The chosen 8mm kernel presents a reasonable tradeoff and is in line with recommendations on how to perform VBM studies (22). Considering that tumor beds and the surrounding tissues show morphological changes between MRI scans not necessarily related to the applied radiation, regions covered by the planning target volume (PTV) were censored during the analysis. The PTV for grade II consisted of the enhanced area on T2-FLAIR with a margin of 1.2 cm (CTV+PTV margins); for grade III-IV it was the gadolinium contrast enhancing tumor on T1-weighted MRI with a margin of 2.2 cm (CTV+PTV margins). This step prevents errors due to tissue misclassification at the tumor bed and in the surrounding tissues, and makes sure that the observed morphological changes are related to the applied radiation.

### Deformation-based morphometry (DBM)

In DBM, the entire brain is analyzed, without pre-specifying the underlying tissue type. During non-linear registration of the individual brains to the standard template, different transformations are applied in each individual (11, 12). This results in 3D deformation fields, in which the local volume changes (expansions or contractions) are described by the Jacobian determinants. This way, the local volumes from each scan are retained in MNI space. By comparing the Jacobian determinants of the pre-RT and post-RT scans, relative volumetric changes after RT were determined for each voxel (**Figure 1A**). Then, these changes were related to the applied dose in EQD2.

**Figure 1.**
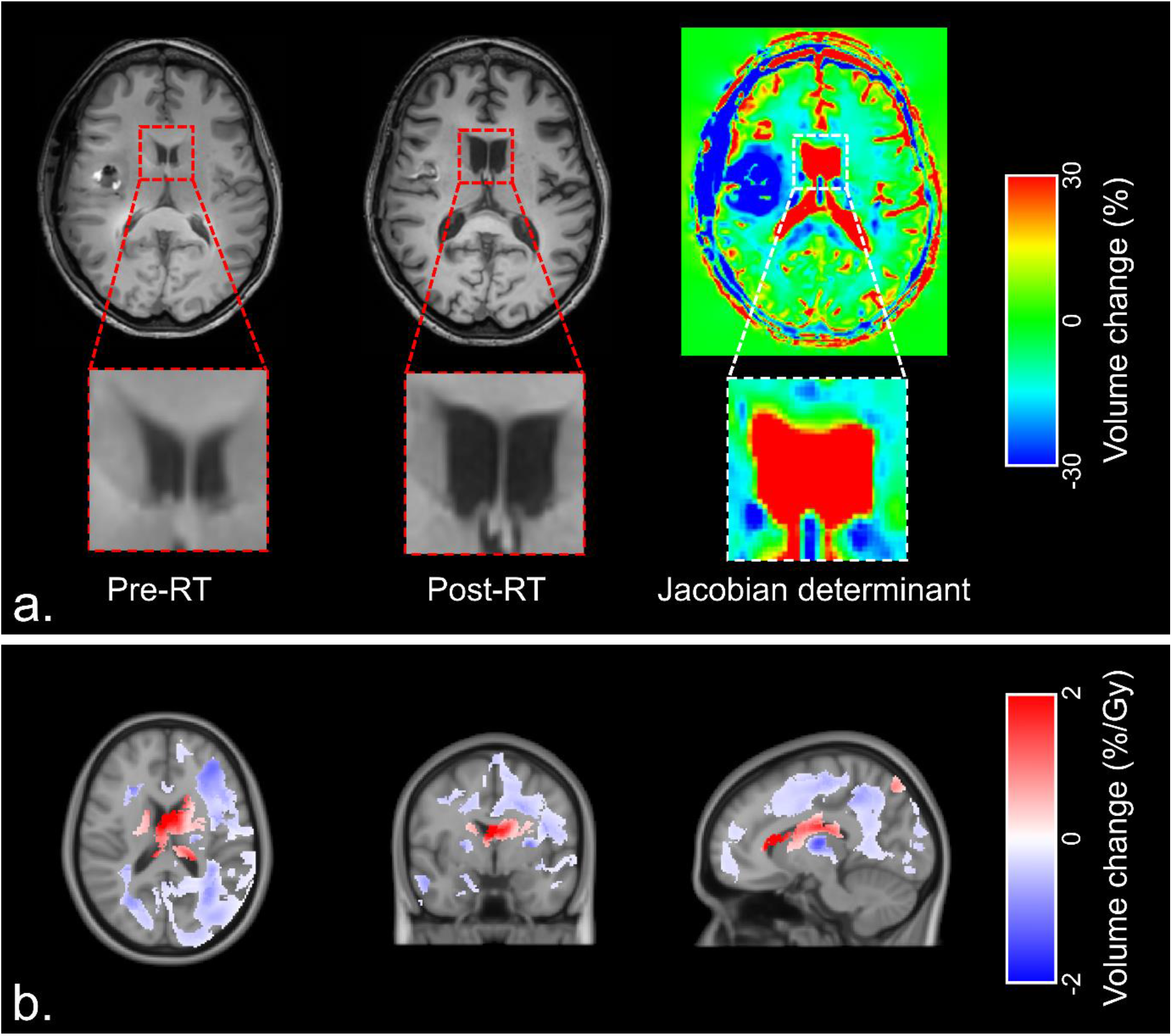
**A**. Example of deformation-based morphometry (DBM), in which two T1-weighted MR (before and after RT) are used to obtain Jacobian determinants. **B**. Areas with significant relation between dose and volume change after 1 year in all patients. Blue indicates local volume loss with increasing radiation dose, red indicates volume increase.

### Voxel-based morphometry (VBM)

In contrast to DBM, VBM is a tissue-specific analysis (15). On both pre-RT and post-RT MRI scans, GM was automatically segmented as part of the processing pipeline (**Figure 2A**). During VBM, the Jacobian determinants from the GM segmentations are used to investigate the modulated (volume preserved) GM changes. Relative changes in volume between the pre-RT and post-RT scans are obtained by comparing Jacobian determinants from the pre-RT and post-RT images, and then correlated to the associated MNI-warped dose maps for every patient.

**Figure 2.**
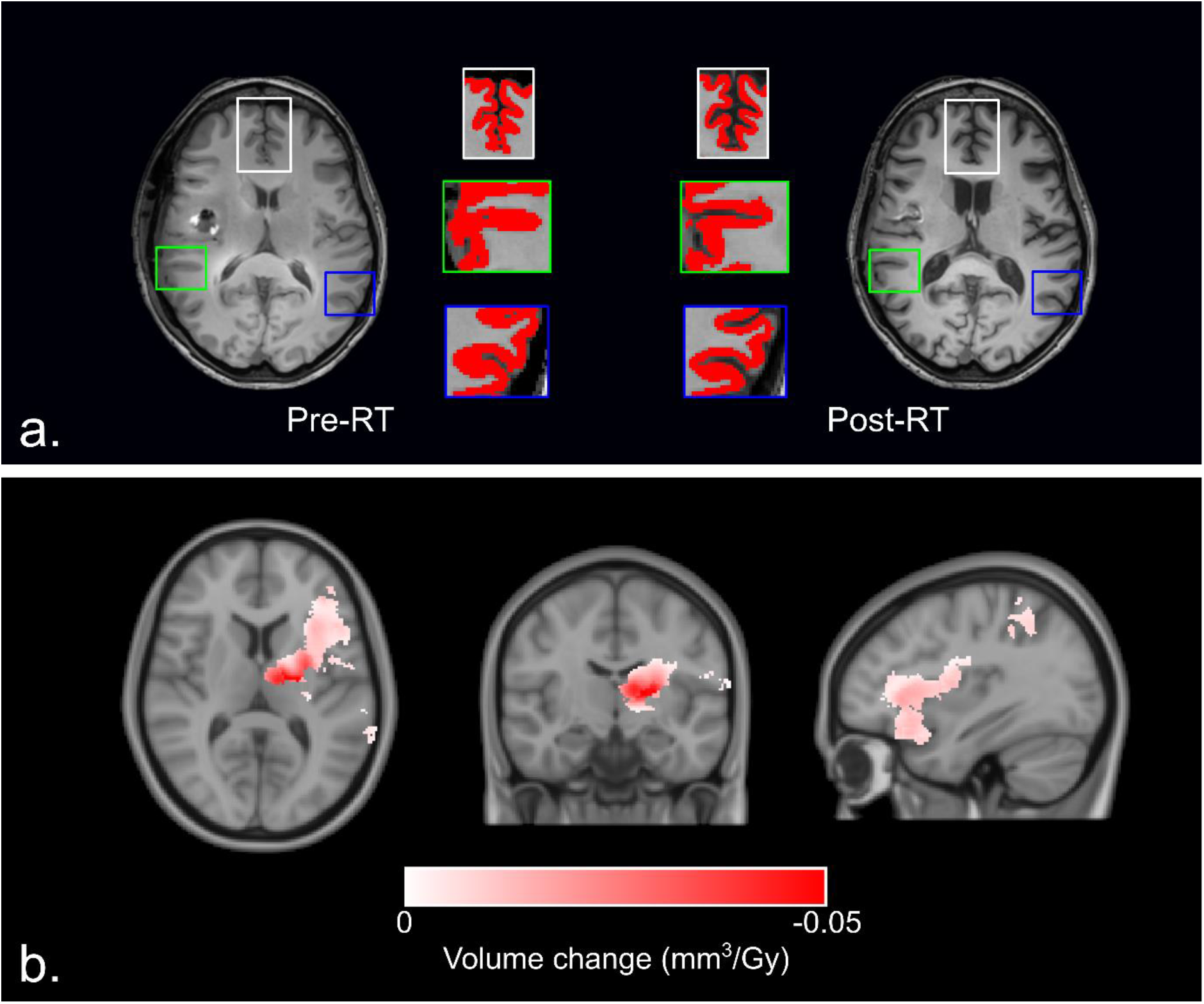
**A**. Example of GM (red mask) changes before and after RT as seen on T1-weighted MRI. **B**. Areas of significant relation between dose and volume change in all patients, as seen with voxel-based morphometry.

### Statistical analysis

Voxelwise and deformation-based statistical comparisons were carried out with a permutation test with 10,000 iterations performed with the permutation analysis of linear models (PALM) toolbox in Matlab (23–25). Significance of a correlation was determined at p_corr_ < 0.05 using family-wise error rate (FWER) adjustment to correct for multiple comparisons, and 3D Threshold-Free Cluster Enhancement (TFCE) to boost the statistical power (26). Age at the time of the diagnosis and sex of the patients were included as nuisance regressors. Tail approximation was used for faster calculations (27). Significant DBM changes were expressed as relative volume changes per received radiation dose (%/Gy), while significant VBM changes are expressed in mm^3^/Gy. In order to report the brain regions in which dose-dependent volume loss occurred, we used the Neuromorphometrics brain atlas (Neuromorphometrics Inc., Somerville, Massachusetts, USA), part of the CAT12 toolbox (20). This atlas divides the brain into 142 regions (of which 126 are GM), based on anatomy and function. The atlas includes all white matter, cerebral cortex, subcortical nuclei, and the ventricles, and allows us to identify the areas that are affected by radiation.

Differences in pre-RT and post-RT volume of GM, WM, CSF, and TIV were examined with a paired Wilcoxon signed rank test, with p < 0.05 as the threshold of statistical significance. This was also done for the ratio between total brain tissue volume (GM+WM) and TIV.

## Results

### Participants

Of the 170 patients who underwent RT for glioma between in 2016 and 2017, 28 fulfilled our inclusion criteria and were selected for further analysis (**Supplementary Figure S1**). Median age at baseline was 51 years, and 61% of patients had a high-grade glioma (**Table 1**). All patients were treated with volumetric modulated arc therapy (VMAT).

**Supplementary figure S1.**
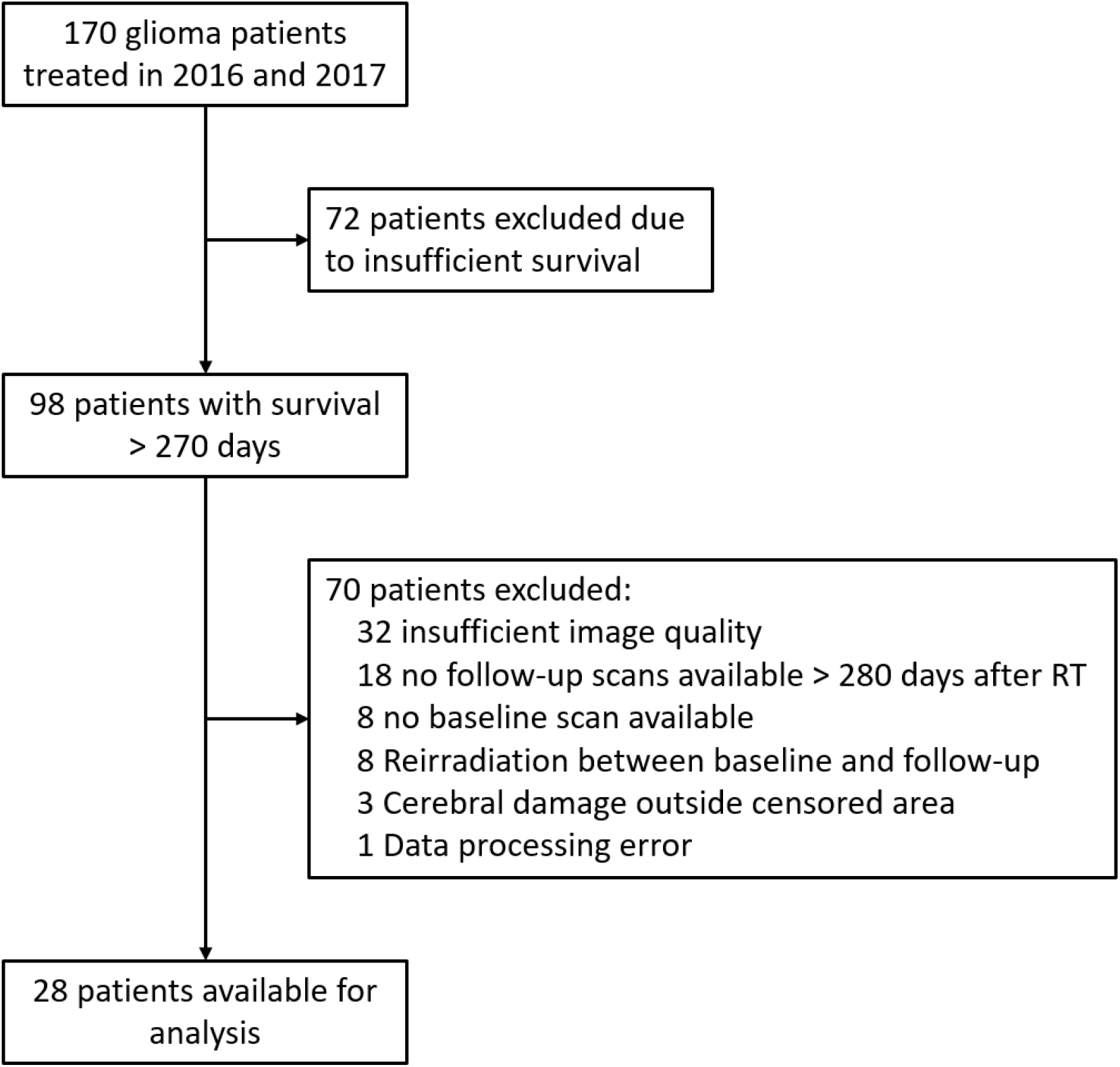
Flow-chart of patient inclusion

**Table 1.** Baseline characteristics of included patients

|  | **N (total n = 28)** |
| --- | --- |
| **Age (median; IQR)** | 51 (38 - 61) |
| **Sex** |  |
| Male | 17 (60.7%) |
| Female | 11 (39.3%) |
| **WHO grade** |  |
| II | 11 (39.3%) |
| III | 7 (25%) |
| IV | 10 (35.7%) |
| **Tumor type** |  |
| Astrocytoma | 13 (46.4%) |
| Oligodendroglioma | 3 (10.7%) |
| Mixed | 1 (3.6%) |
| Ganglioglioma | 1 (3.6%) |
| Glioblastoma | 10 (35.7%) |
| **Prescribed dose** |  |
| 28 × 1.8 = 50.4 Gy | 11 (39.3%) |
| 30 × 1.8 = 54 Gy | 2 (7.1%) |
| 30 × 20 = 60 Gy | 15 (53.6%) |
| **Chemotherapy** |  |
| None | 4 (14.3%) |
| Temozolomide | 20 (71.4%) |
| PCV | 4 (14.3%) |
| PCV = procarbazine, lomustine and vincristine |  |

### DBM

Areas of significant volume reduction with increasing dose 1 year after RT were seen throughout the brain as clusters of affected voxels (**Figure 1B**). This loss of brain tissue consequently led to an increase in ventricle volume, which can be seen as clusters of voxels showing volume increase. Applying the Neuromorphometrics brain atlas revealed that, out of a total of 142 brain regions, 104 (73.2%) contained voxel clusters showing dose-dependent volume loss. Both WM and GM were affected, and within the latter both cerebral cortex and subcortical nuclei show volume loss. Volume loss rates ranging from at least 5.3 to maximal 15.3%/30 Gy were seen in the cerebral cortical regions in which more than 40% of voxels were affected (**Supplementary Table 1**). In the subcortical GM the bilateral hippocampus, thalamus, putamen and globus pallidus show volume loss (**Table 2**) of at least 6.0% and maximal 17.5% per 30 Gy. The left and right cerebral white matter contained clusters both showing loss rates of 11.7%/30 Gy. Conversely, a significant lateral ventricular volume increase was observed, with a mean rate of 52.1 %/30 Gy for the left and right lateral ventricle, respectively. Complete results of DBM analysis are presented in **Supplementary Table 2**.

**Supplementary table 1.** Cortical regions in which more than 40% of voxels show dose-dependent volume loss in deformation-based morphometry

| Region name | Affected voxels<br>within area (%) | Relative volume<br>change (%/30 Gy) | p-value |
| --- | --- | --- | --- |
| Left Frontal Operculum | 67.7 | 15.0 | <0.01 |
| Left Anterior Insula | 51.6 | 10.5 | <0.01 |
| Left Superior Temporal Gyrus | 51.4 | 5.3 | <0.01 |
| Left Middle Occipital Gyrus | 46.7 | 10.8 | <0.01 |
| Left Inferior Frontal Angular Gyrus | 45.1 | 15.3 | <0.01 |
| Left Supplementary Motor Cortex | 44.5 | 9.3 | <0.01 |
| Left Middle Temporal Gyrus | 43.9 | 8.4 | <0.01 |
| Left Planum Temporale | 43.8 | 7.0 | <0.01 |
| Left Inferior Occipital Gyrus | 41.4 | 10.5 | <0.01 |
| Left Precuneus | 40.0 | 10.3 | <0.01 |

**Table 2.** Significant dose-dependent changes in subcortical nuclei after 1 year, as shown with deformation-based morphometry

|  |  | Affected voxels<br>within area (%) | Relative volume<br>change (%/30 Gy) | p-value |
| --- | --- | --- | --- | --- |
| Amygdala | Left | 0 | - | - |
|  | Right | 20.8 | 11.8 | <0.01 |
| Caudate nucleus | Left | 0 | - | - |
|  | Right | 1.1 | 8.6 | 0.02 |
| Globus pallidus | Left | 1.3 | 8.8 | <0.01 |
|  | Right | 14.6 | 10.0 | 0.02 |
| Hippocampus | Left | 21.5 | 6.0 | <0.01 |
|  | Right | 4.3 | 8.0 | <0.01 |
| Nucleus accumbens | Left | 0 | - | - |
|  | Right | 0 | - | - |
| Putamen | Left | 18.6 | 16.9 | <0.01 |
|  | Right | 21.8 | 12.7 | 0.02 |
| Thalamus | Left | 40.5 | 17.5 | <0.01 |
|  | Right | 2.8 | 12.0 | <0.01 |

### VBM

Dose-dependent changes in cortical and subcortical GM volumes were observed in 42 (33.3%) of the 126 brain atlas GM regions. Significant volume loss with increasing dose was again seen both in the cortical and subcortical GM (**Figure 2B**), largely as clusters of affected voxels within the left hemisphere. Of the subcortical nuclei, the left thalamus, caudate nucleus, globus pallidus and putamen contained clusters of dose-dependent volume reduction. In the seven regions in which more than 25% of voxels showed volume decrease, changes between 5.0% and 21.2% per 30 Gy were seen (**Table 3**). No regions showed dose-dependent increase in GM volume after RT. Complete results of VBM analysis are presented in **Supplementary Table 3**.

**Table 3.**
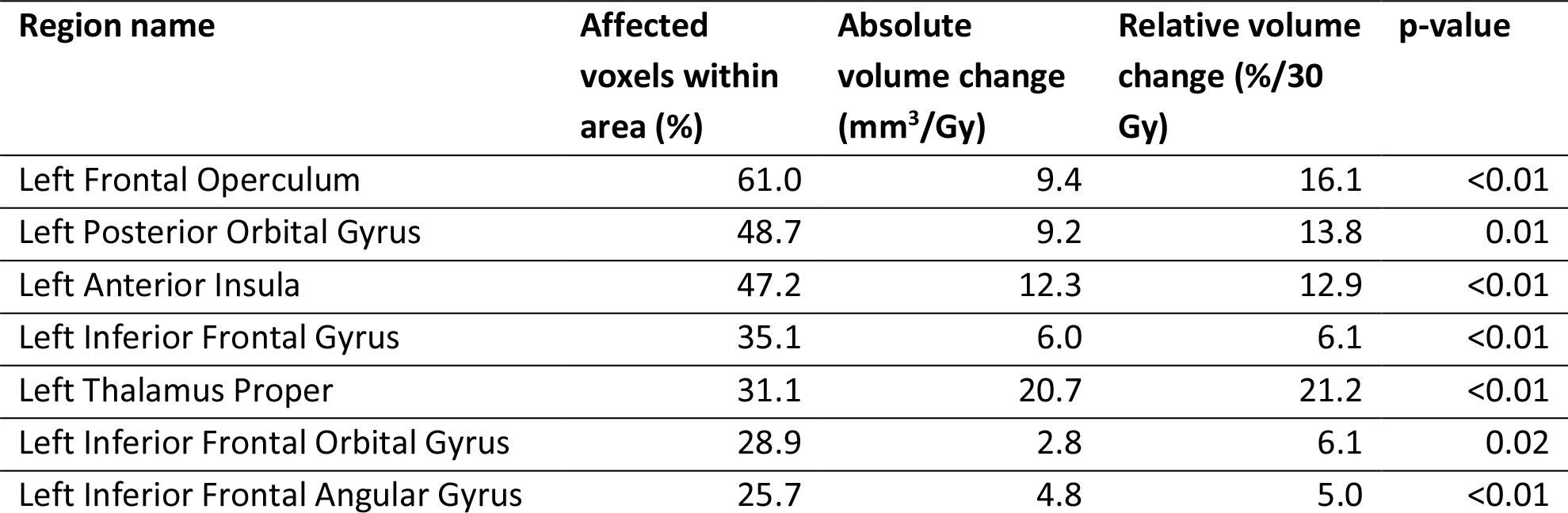
Brain regions in which more than 25% of voxels show volume decrease after one year, as shown with voxel-based morphometry

### Total brain volumes

Median total brain volume (excluding the ventricles) before RT was 1131.9 cc, with a significant median decrease by 43.2 cc (3.8%) 1 year after RT. Mean size and relative post-RT changes for each volume are shown in the **Table 4**. Significant changes are seen in all brain regions, with GM and WM showing decrease of 2.1% and 5.8%, respectively. Expectedly, CSF showed an increase in volume. The total intracranial volume did not change between the two MRI scans. Additionally, the ratio of total brain tissue volume and TIV changed from a median of 80.0% to 75.7% (p=0.03) after one year.

**Table 4.** Changes in brain tissue volumes after RT

| Region | Median pre-RT volume (cc) | Median volume difference (cc) | 95% CI of difference | p |
| --- | --- | --- | --- | --- |
| TIV | 1386.1 | 2.2 | -11.0 – 17.6 | 0.81 |
| GM | 608.2 | -12.8 | -24.8 – -4.7 | < **0.01** |
| WM | 520.6 | -30.0 | -42.0 – -20.1 | < **0.01** |
| CSF | 288.0 | 44.3 | 21.3 – 75.5 | < **0.01** |

CSF = Cerebrospinal fluid; GM = Grey matter; TIV = Total intracranial volume; WM = White matter

## Discussion

We used pre-RT and post-RT MRI scans from glioma patients to assess intracranial morphological changes in the brain after fractionated photon radiotherapy. In addition to reports on specific susceptible areas (3–10), we found that the entire brain is susceptible to radiation induces morphological changes after RT for brain tumors. Total grey matter volume and total white matter volume were reduced by respectively 2.1% and 5.8%, with a compensatory increase in CSF volume. The observed rate of total brain volume change of 3.8% (combined GM and WM) is ten times higher than the normal annual atrophy rate of 0.33% (28). Deformation-based analysis of the entire brain showed volume reductions in white matter, cerebral cortex, and subcortical grey matter. The GM was further analyzed with a voxel-based analysis, again showing susceptibility in a third of cortical regions and subcortical nuclei. Finding volume changes in three distinct types of analyses strengthens the evidence for a whole brain volume-reducing effect of radiotherapy.

This is not the first study looking into the morphological changes seen after RT for brain tumors. The group of Karunami et al. and Seibert et al.(7) were the first to show cortical susceptibility to dose-dependent thinning. Our recent study on this phenomenon confirmed these findings, and identified three cortical areas of heightened susceptibility, showing thinning rates after 30 Gy comparable to aging by a decade (5). Subcortical structures such as hippocampus, amygdala, have also been shown to be vulnerable to volume changes after RT (9, 10). We have repeated these analyses, and observed comparable results (8). Furthermore, we found that in addition to these two structures, the thalamus, globus pallidus, nucleus accumbens, and putamen also show an association between radiotherapy dose and post RT volume loss.

Connections have also been made between these morphological changes and observed cognitive decline, especially for hippocampal volume loss. Gondi et al. (29) showed an association between radiation dose and memory impairments, specifically regarding delayed recall performance. The effect of radiotherapy with or without hippocampal avoidance was further studied in a phase III trial (30). Similar overall and progression-free survival were observed, but with lower risk of cognitive failure and better preservation of executive function, learning and memory.

Most of the abovementioned studies have investigated a specific part of the brain. However, the brain is a complex network of interconnected brain regions (31). This is especially crucial when considering higher order cortical functions like cognition, which have been shown to rely on large-scale neural networks (32). This means that analysis of only cerebral cortex or subcortical structures results in an incomplete picture of possible substrates of post-RT cognitive decline. We have therefore used DBM to analyze the entire brain (WM, cortex and subcortical nuclei), without prior specification of tissue type or location. This resulted in the finding that local susceptibility to radiation-induced damage is present throughout the brain. We conclude from this that a holistic approach to the discovery of etiology and possible prevention of cognitive decline is called for. We need to know the relation between dose and cognitive outcomes for the entire, not just for selected regions. Considering the fact that avoiding a structure like hippocampus leads to increased photon doses being delivered to other areas of the brain (33), we need to be sure these regions don’t have a similar susceptibility to radiation damage.

Because we do not have neurocognitive data on the studied patients, we have to look at morphological changes in other brain diseases to understand the implications of our results. Especially in Alzheimer’s disease, numerous associations have been found between the volumes of cortical and subcortical GM and the development of cognitive impairments (34–38). Additionally, the volumetric change in at least one GM structure has been linked to cognitive disabilities in normal ageing (39), Parkinson’s disease (40–43), Huntington disease (44), and multiple sclerosis (45).

These results may make us reconsider the currently used organs at risk (OAR) in radiation treatment of brain tumors. Several institutions have implemented hippocampal avoidance whole-brain RT to prevent cognitive decline in patients with brain metastases. However, during treatment plan optimization, higher doses are being delivered to surrounding tissue when lowering dose in a specific structure (33). We have found that this surrounding tissue can be similarly susceptible to radiation-induced damage, which warrants future research into the relation between morphologic changes in the entire brain and cognitive outcomes. Then it can be conclusively deducted which parts of the should be considered OARs, and therefore should receive as little dose as possible, to limit or even prevent radiation-induced cognitive decline. Modern techniques such as VMAT and intensity modulated proton therapy (IMPT) could help us to specifically spare healthy brain tissue, should a relation between dose and cognition be found (46, 47). Avoiding critical brain structures could lead to better preservation of cognition, and would therefore improve quality of life in brain tumor patients after treatment.

The biggest limitation to our study is the limited sample size. Due to the requirement of high quality T1 MRI scans before and after RT, only a small portion of scans were eligible for inclusion. We were therefore only able to study a linear effect between the two time points. The sample size could also be the cause of the significant results of VBM to be predominantly within the left hemisphere. However, this meticulousness improves the reliability of our results, as they are unlikely to be affected by image quality.

Secondly, RT was not the only treatment received by patients in our cohort. Most also received previous surgery and concurrent or adjuvant chemotherapy, meaning that these treatments could also have affected the observed morphological changes. However, the baseline scans used for analysis were made after surgery, which means surgery is less likely to have an effect on the outcome. Chemotherapy, which has been shown to cause changes in brain tissue in non-neurological malignancies (48, 49), could have had a diffuse effect on brain tissue volumes. As we related the locally applied RT dose to brain morphology, we expect the role of chemotherapy to be limited.

Finally, we do not have prospectively registered neurocognitive data on the patients in our cohort. This means we cannot conclude on clinical implications of the observed morphological changes, and therefore we cannot give strong recommendations to alter current RT strategies.

## Conclusion

To conclude, we have found that radiotherapy dose is associated with morphological changes in the entire brain. Furthermore, these changes are linked to increased dose. This may lead us to consider revising the current avoidance strategies, as now only a limited number of areas are considered organs at risk, while our data suggests the whole brain volume should be taken into account. Before this can be done, more data on the relation between these morphological changes and cognitive and other neurological outcomes after radiotherapy are needed.

## Data Availability

As we have not obtained informed consent to share patient data, we cannot make our clinical data available.

## Notes

**Conflict of interest** Nothing to declare

### Competing Interest Statement

The authors have declared no competing interest.

### Funding Statement

Nothing to declare

### Author Declarations

Ethical oversight, including the need for informed consent, for this retrospective study was waived by our institutional review board (Medisch Ethische Toetsingscommissie Utrecht), with reference number 18-274.

